# Limitations of estimating antibiotic resistance using German hospital consumption data - A comprehensive computational analysis

**DOI:** 10.1101/2024.05.06.24306930

**Authors:** Michael Rank, Anna Kather, Dominik Wilke, Michaela Steib-Bauert, Winfried V. Kern, Ingo Röder, Katja de With

## Abstract

For almost a century, antibiotics have played an important role in the treatment of infectious diseases. However, the efficacy of these very drugs is now threatened by the development of resistances, which pose major challenges to medical professionals and decision-makers. Thereby, the consumption of antibiotics in hospitals is an important driver that can be targeted directly. To illuminate the relation between consumption and resistance depicts a very important step in this procedure. With the help of comprehensive ecological and clinical data, we applied a variety of different computational approaches ranging from classical linear regression to artificial neural networks to analyze antibiotic resistance in Germany.

These mathematical and statistical models demonstrate that the amount and particularly the structure of currently available data sets lead to contradictory results and do, therefore, not allow for profound conclusions. More effort and attention on both data collection and distribution is necessary to overcome this problem. In particular, our results suggest that at least monthly or quarterly antibiotic use and resistance data at the department and ward level for each hospital (including application route and type of specimen) are needed to reliably determine the extent to which antibiotic consumption influences resistance development.

## Introduction

The global increase in antimicrobial resistance (AMR) is one of the greatest challenges facing medicine today. A lot of previous publications have estimated the effect of AMR on deaths including reports with up to 10 million deaths annually by 2050 [1]. There is sufficient evidence that antibiotic consumption is correlated with the development of resistance [2]. Even more, if no global countermeasures are taken, simple infections caused by resistant, especially gram-negative, pathogens will no longer be treatable in the future [3].

In order to counteract the development of antibiotic resistance, so called Antimicrobial Stewardship (AMS) programs are implemented worldwide. These programs encourage the rational use of antibiotics in order to improve patient outcomes, ensure cost-effective therapy, and reduce adverse sequelae of antimicrobial use. Systematic reviews have shown that interventions to decrease excessive prescribing were associated with reduction in *Clostridioides difficile* infections and colonization or infection with resistant gram-negative bacteria, methicillin-resistant *Staphylococcus aureus* and vancomycin-resistant *Enterococcus spp*. [4–6]. Therefore, international and national guidelines on AMS recommend the reduction of certain substances/classes to control the incidence of infection with resistant bacteria and *Clostridioides difficile* [7–10].

An important prerequisite for guideline-based AMS measures is the collection of resistance and antibiotic consumption data. The AMS-guideline by the German Society for Infectious Diseases [7] recommends collecting the data on different levels (e.g. patient, ward, hospital), depending on the aim of the intervention. The dynamics between exposure (antibiotic use) and effect (resistance) are also important for the interpretation of many studies. In particular, on the one hand, restricting the use of antibiotics is usually only effective with a delay of several months (∼6) with regard to the development of resistance. On the other hand, the effects of the interventions on the use of the substance are measurable after just a few weeks [4]. Therefore, the delayed effect of changes in antibiotic use on the development of resistance should always be taken into account.

It has been shown that mathematical and statistical models are a useful tools to investigate the extent to which antibiotic use influences resistance development, even if accurate data on patient level are not available as in Germany [11]. Therefore, in this study, we have implemented different computational approaches, ranging from linear regression models to artificial neural networks to determine the extent to which antibiotic use influences resistance development. Furthermore, we use our analysis to investigate current limitation induced by restrictions of data availability and to identify data collection strategies that could overcome these problems. To the best of our knowledge, this is the first study to use such in-depth model development to investigate the relationship between consumption and resistance using nationwide data from Germany.

## Methods

For the present work we used quarterly consumption and resistance data over a 5-year period (2015-2019). We obtained antibiotic consumption data on department, specialty and ward level from a Germany-wide antibiotic surveillance project, called ADKA-if-DGI-project [12], a cooperation between the German Hospital Pharmacists Association (ADKA), the Division of Infectious Diseases of the university hospital Freiburg (if) and the German Society of Infectious Diseases (DGI) including more than 300 acute care hospitals. Consumption data on antibacterial drugs are expressed as RDD per 100 patient days [13,14]. Furthermore, we used antibiotic resistance data for *E. coli* from the Antibiotika-Resistenz-Surveillance (ARS)-database [15] provided by the Robert Koch-Institute (RKI). The ARS database collects routine data on antibiotic susceptibility testing of all clinical pathogens and sample types from ∼80 participating laboratories which supply ∼600 hospitals. Routine laboratory testing data are reported as proportions of R (resistant), I (intermediate) and S (sensitive) strains according to national quality standards (MIQ, Mikrobiologisch-infektiologische Qualitätsstandards [16]). Resistance data are only provided at the federal state level and are not available for each individual participating hospital. If the number of isolates to be tested is less than 50, no information on the proportion of resistant pathogens is given. The average number of isolates of some federal states is low, so we decided to aggregate them into larger regions (Thuringia & Saxony-Anhalt, Brandenburg & Mecklenburg-Western Pomerania, Schleswig-Holstein & Hamburg, Lower Saxony & Bremen, Rhineland-Palatinate & Saarland) to increase data availability. To evaluate the results, in-house consumption and resistance data from the university hospital Carl Gustav Carus in Dresden (UKD) had been used with the same structure. Hence, no information about individual patients could and can be identified in all used data sets.

Ethical approval was not required, because the project was based on epidemiological data. Research involving human subjects, human material and specific human or personalised data was not carried out. All data was anonymised regarding the hospital names for antibiotic consumption. Antibiotic resistance data was collected by the ARS database on federal state level and does not include any personal information either.

Firstly, we used descriptive statistics and visualizations to determine some possible patterns and trends in the data. Secondly, we applied computational models in order to relate the antibiotic use and development of resistance and to uncover possible interactions. Therefore, we considered and compared a wide variety of methods with the goal of obtaining robust results as different approaches focus on different characteristics of the data.

In all models, corresponding hospital antibiotic consumption data from each region were aggregated to match the resistance data. We are working with *recommended daily doses* (*RDD*) instead of *defined daily doses* (*DDD*) values for accuracy reasons, as they match the actually prescribed dosages more precisely [17,18]. As mentioned before, the proportions of resistant pathogens are derived by different sample types. Due to the low number of blood samples, we used all sample types to obtain the target values for the models. We also included various quarterly time shifts between consumption and resulting resistance in all presented models. We considered both a classical multiple linear regression and a linear mixed effect model, which complements the fixed effects with additional random effects, accounting for dependency among data points. Within the mixed-effect model, the regions served as random effects, because we are not interested in the effect of a particular region, but in the regional effect as a general source of heterogeneity. Furthermore, we used departments (internal vs surgery), the distinction between general ward and ICU, and the consumption in RDD per 100 patient days of some selected antimicrobial substances as fixed effects. Additionally, we incorporated the number of patient-days as well as time in years and quarters into the model to account for potential confounding effects, where patient-days refer to the total number of days each patient occupies a hospital bed. We utilized the proportion of *E. coli* isolates being reported resistant to ciprofloxacin or cefotaxime as the dependent response variable. For each of the two substances we fitted a separate model. All linear regression and mixed-effect models were calculated using the statistical software R [19]. To determine the antibiotics used as predictors, we firstly included ciprofloxacin and cefotaxime in the according models as important representatives of the antibiotic class of fluoroquinolones and third generation cephalosporins respectively. Secondly, based on similar biochemical functionality, we added all substances of the corresponding antibiotic class that were used during the considered period. Thirdly, based on a successive model selection using the AIC criterion [20], we added or removed substances as predictors and potentially transformed them according to their distributions to generate some possible regression model configurations. Thereby, substances of all antibiotic classes (e.g. penicillins, cephalosporins etc.) were possible candidates. To check for and to avoid overfitting artefacts, we split the data randomly into a *training* (90%) and *test* (10%) data set. A detailed justification for this is given in the following paragraph. Moreover, we fitted the regression models to the training data and, thereafter, applied them to the test data in order to evaluate and compare the accuracy. Based on the assumption that there should be no resistance of *E. coli* to the two investigated substances without any antibiotic consumption, we assumed a *zero intercept* (i.e. regression line to pass through the origin).

In addition to this classical statistical modelling framework, we applied several further *machine learning* methods to our problem. In terms of performance, *artificial neural networks* (i.e., *deep learning methods*) prevailed over other methods, which were *random forests* (RF) and *support vector machines* (*SVM*) [21]. In the following, we will, therefore, limit ourselves to the description of the artificial neural networks that we implemented in Python with the help of the *tensorflow* and *keras* package [22]

We used the consumption of all antibiotic substances as the input and the *E. coli*-resistance to ciprofloxacin and cefotaxime as the output, both of which are fitted in the same model. Additionally, we incorporated the information about the respective regions, departments, and general ward / ICU into the model as *Dummy variables* using so-called *one-hot encoding* [23], where categorical variables are transformed into several binary vectors. We applied a keras hyperparameter optimization framework to obtain a model containing three hidden layers with 50, 50 and 30 neurons and *ReLU* activation function [24]. *Adam*, an adaptive stochastic gradient descent method [25], served as the optimizer. To improve the quality of the results, we also incorporated the concept of *zero intercept* in linear models (see above). Technically, this was done by adding a fixed number of zero lines to the data. In order to compare the different models to each other, we used the *mean squared error* (*MSE)*. As artificial neural networks require a lot of data to be trained properly, we decided for the split into 90% training and 10% test data mentioned above.

In order to evaluate the in-house data of the UKD on ciprofloxacin and cefotaxime resistance of *E. coli*, we used linear models only, because neural networks do not lead to meaningful results because of the small sample size. Starting with the same models as for the main data described above, we removed the random effects, as there are no regional differences. Furthermore, we performed a model selection in the same way and used the coefficient of determination R^2^ to compare this linear model with the regional linear model, as both models were trained on different datasets.

## Results

Antibiotic surveillance data from the ADKA-if-DGI-project show that overall antibiotic use has remained stable over the five-year period (2015-2019), see Figure 1a.

**Figure 1:**
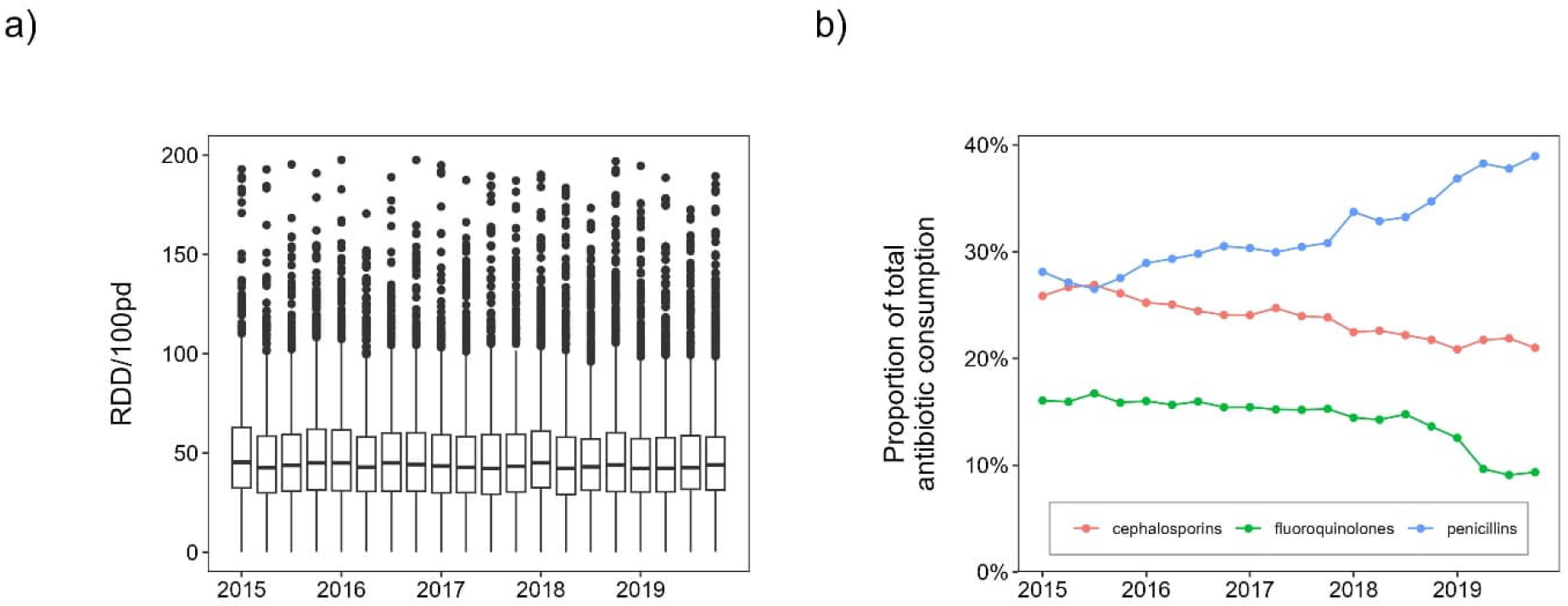
Time course of antibiotic consumption. a) Boxplots of total antibiotic consumption of all individual hospital departments and ward types for each quarter from 2015 to 2019. b) Average proportion of total antibiotic consumption for each quarter and some selected antibiotic classes.

Beta-lactams and fluoroquinolones accounted for about 74% of total consumption in 2019. However, the composition of antibiotic substances has changed over time. On average across all hospitals, fluoroquinolone use decreased ∼6% and cephalosporin use decreased ∼4% from 2015 to 2019, while penicillin use increased ∼10%, see Figure 1b for corresponding trends over time. Figure 2a and b show boxplots of the proportion of *E. coli* resistant to ciprofloxacin and cefotaxime in German hospitals (ARS database, all sample types). We can observe a nonlinear curve of median ciprofloxacin resistance-levels with peaks in the first quarter of 2015 and second quarter of 2017 (both ∼20%) and the lowest value at the end of 2019 (∼17%) with an overall decreasing trend. There is no clear trend in cefotaxime resistance in *E. coli* and values fluctuate around 12%. Similar consumption patterns emerge for the UKD, with an increase of penicillins (+15%) and a decrease of fluoroquinolones (−10%) and cephalosporins (−1%). Beta-lactams account for 66% (penicillins alone for 41%) and fluoroquinolones 9% of total antibiotic consumption in 2019. In-house UKD *E. coli*-resistance to ciprofloxacin decreased from 27% in 2015 to 16% in 2019, while *E. coli*-resistance to cefotaxime decreased from 16% to 13%. Note, however, that the median values for individual quarters vary considerably, which is not the case with the nationwide ARS-RKI data.

**Figure 2:**
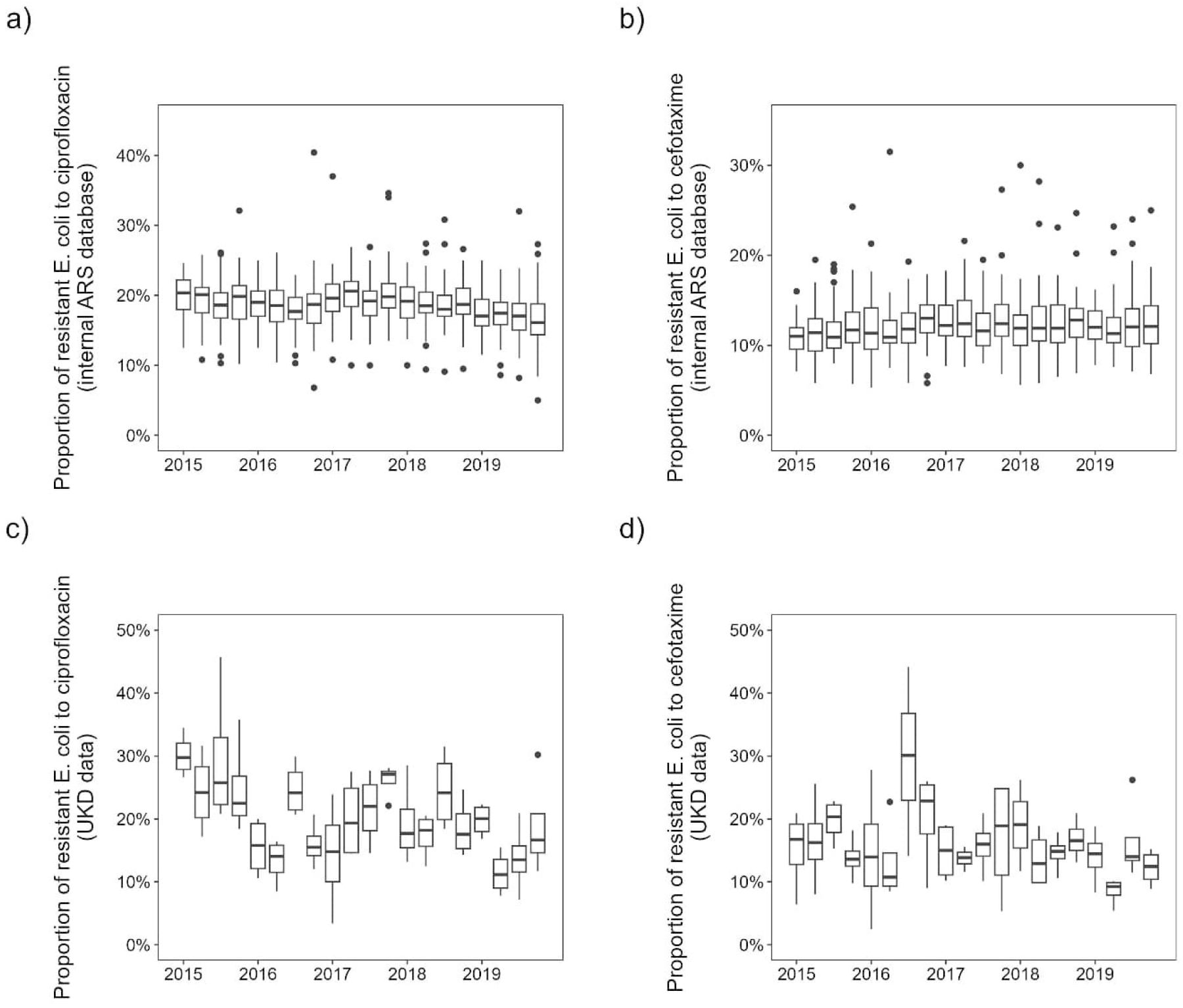
Boxplots of the proportion of resistant *E. coli* to. a) ciprofloxacin and b) cefotaxime of all regions for each quarter from 2015 to 2019 for German hospital department and ward types in internal ARS database. The boxplots in c) and d) show the results for the in-house UKD department and ward type data accordingly.

Both the linear models and the artificial neural networks described in the Methods section showed a sufficiently good model fit (in terms of MSE), and the respective model assumptions of the linear models are adequately met, justifying their application. We were able to improve the predictions of the linear model by introducing the zero intercept. There were no major differences in the models due to different time shifts between antibiotic use and resistance. Note that for future studies one or even several time shifts might be crucial, as mentioned in the introduction.

Although the fitting itself worked well (in terms of minimizing MSE), the results of the different methods are very inconsistent and lead to conflicting interpretations. For instance, there are discrepancies in the predicted impact of consumption of certain antibiotics on *E. coli*-resistance. This effect occurred not only between different methods, but also within each individual model, as adding or neglecting a single predictor partially resulted in very different model parameters. Thus, none of our models is stable against small deviations. To assess the performance of these regional models at the hospital level, we applied them to our available UKD consumption and resistance data. Ideally, we would expect an error in the order of magnitude of the regional data level. However, very high MSE occurred in both models-types, indicating that the models trained on the aggregated regional data do not perform well at the individual hospital level. Since these discrepancies occurred for different model types and all quarterly time shifts, we conclude that the reason for this is to be found in the data rather than the methodology. Specifically, we suppose the resistance data to be the major reason for this problem, because it is provided only at the federal state level but collected at the clinical level. Therefore, relevant effects are averaged out and the data become too homogeneous. This results in quarterly values that are very close, while in some quarters there may have been large differences in both antibiotic consumption and resistance over time that are not apparent in aggregated form. This becomes clear when comparing the variance of *E. coli-*resistance to ciprofloxacin of the UKD data set with the variance of the Saxon data of the ARS RKI data set: With a variance of 54.6, it is significantly higher than in the Saxon data with a value of 8.2. The values for cefotaxime are correspondingly 46.6 and 8.2. This can also be seen in Figure 2, the differences in the means of the UKD data are much larger than in the ARS RKI data. The higher scatter in the UKD data for both substances is likely due to the fact that we are comparing consumption and resistance values from a single hospital to data aggregated across more than twenty hospitals in the entire federal state. Therefore, we conclude that individual data points, i.e. the quarterly reported antibiotic resistance levels, are too similar to each other in the ARS RKI data. This is a direct result from aggregating the data to federal states and leads to the depicted loss of presumably relevant variance components.

To demonstrate the benefit that can be gained from hospital level data, we took a closer look at linear model results using only the in-house UKD *E. coli* ciprofloxacin resistance data. We were able to accomplish an average R^2^-value of approximately 0.93 in contrast to 0.76 for the regional linear model. Note that this analysis is for illustrative purposes only and should not be taken as a definitive result, as looking at a single hospital results in a very small sample size and a potential *overfitting* (i.e. the particular hospital could in principle be an exception to the rule), so the results should be viewed with some caution. However, this highlights the importance of systematically collecting and providing matched antibiotic consumption and corresponding resistance data at the hospital and department level across the country. This applies analogously to the prediction of resistance to ciprofloxacin and is therefore not presented here. It should be noted that the in-house data were utilized to show the possible prospects that can be gained from hospital level data.

## Discussion

The aim of this analysis was to estimate antibiotic resistance using hospital consumption data in Germany, taking as an example the proportion of resistant *E. coli* to the drugs ciprofloxacin and cefotaxime, based on consumption numbers of all antibiotics applying computational models ranging from classical linear models to artificial neural networks. Our study showed that the quality of the data available on antibiotic consumption and resistance in Germany is not sufficient to develop reliable and stable models. For the models trained on resistance data from the RKI and consumption data from the ADKA-if-DGI-project, the model performance on the in-house hospital data was very poor. The main reason for this seems to be the aggregation of resistance data at the federal state level, which is masking some presumably important characteristics, see Results. To the best of our knowledge, this is the first study to use such in-depth model development to investigate the relationship between consumption and resistance development using nationwide data for Germany.

To demonstrate that the developed models perform better when they use a higher resolution of the data sources, i.e. quarterly resistance and consumption data at hospital and department level, we pilot-tested our linear model on data from one hospital, which yielded promising results. This analysis substantiated our hypothesis that aggregating data over (all) hospitals of each region leads to a depicted loss of variance relevant component. That is, if we could match the consumption and resistance data of every hospital instead of just considering regions, we could work with more than 25 times the sample size and gain more variance components. Especially for machine learning algorithms such as the neural networks presented above, this could lead to much better model performance [26]. Depending on the aggregation, the considered exposure of a population to antibiotics cannot ensure that the part of the exposed population corresponds exactly to the part in which resistances are observed [11]. However, the higher the aggregation, the stronger uncontrolled influences become and the more uncertain the causality in the relationship between exposure (antibiotic prescription) and effect (resistance) becomes. In addition, it would be desirable to have data available on a monthly basis to be able to resolve local and temporary events and fluctuations even better, as for instance an outbreak of *MRSA* in a hospital department. However, this concerns both the consumption and resistance data and is associated with a significantly higher effort.

Published surveillance reports in the Netherlands (“Nethmap”) [27] and Denmark (“Danmap”) [28] also only contain annually aggregated data, but on a nationwide basis. Nevertheless, in both countries, antibiotic consumption data are actually available at patient level (Netherlands: electronic medical record) or at hospital level (Denmark: monthly collection by querying sales data from all healthcare providers) for all acute care hospitals. In their surveillance report “Danmap 2020” [29] Denmark reports monthly antimicrobial consumption data for the first time, while it is also mentioned that at some hospitals they have antimicrobial consumption data on patient level via electronic medical record [29]. Resistance data is also available on patient level via their Danish Microbiology Database (MiBa) which records all microbiological test results, but this is not publicly available. Access needs to be approved by the Departments of Clinical Microbiology (DMC) and e.g. the Danish Data Protection Agency [30].

This study clearly shows that more detailed data is needed to determine the extent to which antibiotic use influences resistance development. For this reason our model requires at least the same data quality as in Denmark. If our model were applied to this data, it could serve as a stimulus for a possible analysis of the interactions between consumption and resistance. Hence, it may be possible to uncover previously unknown relationships between the consumption of certain antibiotics and patterns of resistance that have emerged.

In the future, we should have resistance and consumption data available in real time at the patient level. This would allow the development of automated early warning systems for physicians and healthcare professionals. With its help, a more rational use of antibiotics will be promoted, thus minimizing the risk of resistance development. First and foremost, the publication of monthly or quarterly antibiotic consumption and resistance data in Germany or Europe for research should be accelerated in the near future to enable answers to these and similarly important research questions to prevent potential humanitarian disasters.

## Acknowledgements

We would like to thank Mr. Kemmerer from the University Hospital Carl Gustav Carus Dresden, Division Information Technology, supporting us with an automated data retrieval to get resistance data from the ARS homepage. And last but not least, we would like to thank our colleagues at the Institute for Virology, Institute for Medical Microbiology and Hygiene, Medical Faculty Carl Gustav Carus, Technische Universität Dresden to provide us with resistance data for the University Hospital Carl Gustav Carus Dresden.

## Declarations

### Ethical statement

Ethical approval was not required, because the project was based on epidemiological data. Research involving human subjects, human material and specific human or personalised data was not carried out. The manuscript does not contain clinical studies or patient data.

### Data availability statement

The data sets and code used in this publication are available in the Zenodo repository under DOI <u>10.5281/zenodo.10684599</u> (https://doi.org/10.5281/zenodo.10684599).

